# Longitudinal protection following natural SARS-CoV-2 infection and early vaccine responses: insights from a cohort of community based dental health care professionals

**DOI:** 10.1101/2021.02.24.21252368

**Authors:** Adrian M. Shields, Sian E. Faustini, Caroline A. Kristunas, Alex M. Cook, Claire Backhouse, Lynsey Dunbar, Daniel Ebanks, Beena Emmanuel, Eddie Crouch, Annika Kroeger, Josefine Hirschfeld, Praveen Sharma, Razza Jaffery, Sylwia Nowak, Samantha Gee, Mark T. Drayson, Alex G. Richter, Thomas Dietrich, Iain C. Chapple

## Abstract

**Background:** The threshold of protection for anti-SARS-CoV-2 spike glycoprotein antibodies and their longevity are not known. Interpretation of serological results in with respect to international reference material can inform this essential question.

**Methods:** 1,507 West Midlands dental care professionals were recruited into this study in June 2020. Baseline seroprevalence of antibodies directed against the SARS-CoV-2 spike glycoprotein was determined and the cohort was followed longitudinally for 6 months until January/February 2021 through the second wave of the COVID-19 pandemic in the United Kingdom, and commencement of vaccination.

**Results:** Baseline seroprevalence was 16.3% in this cohort, compared to estimates in the general population of between 6-7%. Seropositivity was retained in over 70% of participants at 3 and 6-month follow up and conferred a 74% reduced risk of infection. During follow-up, no PCR-proven infections occurred in individuals with a baseline anti-SARS-CoV-2 IgG level greater than 147.6 IU/ml with respect to the World Health Organization international standard 20-136. Post-vaccination, antibody responses were more rapid and of higher magnitude in individuals with who were seropositive at baseline.

**Conclusion:** Natural infection leads to a serological response that remains detectable in over 70% of individuals 6 months after initial sampling and 9 months from the peak of the first wave of the pandemic. This response is associated with protection from future infection. Even if serological responses wane, a single dose of the Pfizer-BioNTech vaccine is associated with an antibody response indicative of immunological memory.

**Funding:** The Association of Clinical Biochemistry and Laboratory Medicine and The Institute for Global Innovation (IGI) of the University of Birmingham.

## Introduction

Seroepidemiological studies of healthcare workers define occupational risk of exposure to the SARS-CoV-2 virus [1-3] and seropositivity is associated with protection from subsequent infection in high-exposure cohorts [4, 5]. Such studies have guided public health planning, the design of healthcare services, and associated infection prevention protocols to mitigate risk and maintain essential care services during the pandemic. However, an absolute level of antibodies that confers protection for a given period of time remains unknown.

Dental care professionals (DCPs) represent a group of healthcare professionals thought to be at high-risk of exposure to SARS-CoV-2 because they routinely operate within patients ‘aerodigestive tract and undertake aerosol generating procedures (AGPs). The risks of occupational exposure to and infection with SARS-CoV-2 in dental teams remains unclear despite studies evaluating aerosol generation and viral recovery in relevant bodily fluids [6, 7]. Many dental practices across the world closed in March 2020 and, in the United Kingdom, did not re-open until June/July 2020, when level 3 personal protective equipment (PPE) was in sufficient supply to enable resumption of AGPs for those in need of urgent dental care. The impact of this policy on patients remains unclear.

In June 2020, we recruited a cohort of 1,507 community and hospital based DCPs from the West Midlands region of the United Kingdom, following the first wave of the COVID-19 pandemic. The longitudinal follow-up of this cohort through the second wave of the COVID-19 pandemic and following the start of widespread vaccination of healthcare workers, afforded a unique opportunity to study occupational risk factors in DCPs, the durability of serological responses and to compare the early kinetics of serological responses following a single dose of the Pfizer-BioNTech vaccine based on prior exposure to the virus. Furthermore, using World Health Organization (WHO) and National Institute for Biological Standards and Control (NIBSC) international reference material, we were able to define an anti-SARS-CoV-2 spike glycoprotein antibody concentration arising following natural infection associated with protection from reinfection for 6 months.

## Methods

### Study recruitment

Registered general dental practitioners (GDPs) in the West Midlands area, were invited by email to participate in a research study on SARS-CoV-2 antibody status in May 2020. GDPs were encouraged to disseminate this invitation to their wider dental teams. A total of 1,716 individuals registered their interest, of which 1,535 attended the first study visit and provided informed written consent. Twenty-three individuals were excluded when they were found to not work in dentistry. 1,507 participants volunteered a venous blood sample that was suitable for serological analysis at their first study appointment. Study participants also provided occupational and ethnodemographic data. The index of multiple deprivation in participants ‘home postcode were sourced from 2019 UK Ministry of Housing, Communities and Local Government statistics [8]. Study data were collected and managed using REDCap electronic data capture tools hosted at the University of Birmingham [9]. Individuals who were seropositive at baseline were recalled three months later (September 2020) to study the persistence of their antibody response. All participants were recalled six months after providing their baseline sample (January 2021). At the time of manuscript preparation, 873 participants had volunteered a repeat blood sample suitable for serological analysis.

### Serological analysis

Serological analysis was performed using a commercially available, CE marked, IgGAM ELISA that measures the total antibody response (IgG, IgA and IgM simultaneously) against the spike glycoprotein (Product code: MK654, The Binding Site (TBS), Birmingham). Detailed descriptions of this assay including its construction, validation and verification have been published previously [10, 11]. The assay is optimized for seroepidemiological studies and demonstrates 98.3% (95% CI: 96.4-99.4%) specificity and 98.6% sensitivity (95% CI: 92.6-100%) in detecting a serological response to the SARS-CoV-2 spike glycoprotein following PCR-positive, non-hospitalised, mild-to-moderate COVID-19. Internal quality control material demonstrates an inter-assay coefficient of variance of 7.2% at the cutoff. Samples are run at a standard dilution of 1/40. To assess the response of individual immunoglobulin isotypes, the above protocol was modified to employ polyclonal sheep-anti-human HRP-conjugated polyclonal antibodies against IgG (1:16,000) and IgA (1:2000) as secondary antibodies. For these assays, a cutoff ratio of 1.0 relative to the existing TBS cutoff calibrators was determined by plotting the pre-2019 negatives (n=90) in a frequency histogram chart. Once the ratio cutoff was determined from the pre-2019 negatives, a cut-off multiplier of 1.0 and 0.71 was established for IgG and IgA respectively.

### NIBSC and WHO standards

In late 2020, the National Institute for Biological Standards and Control (NIBSC) developed international reference material (IRM) for the purposes of traceability and calibration of SARS-CoV-2 serological tests. These include NIBSC 20/136, the first World Health Organization International Standard for anti-SARS-CoV-2 immunoglobulin [12] and NIBSC 20/162. Serial dilutions of these IRM were run in triplicate on the SARS-CoV-2 IgG assay described above. A receiver operator characteristics curve was constructed using baseline anti-SARS-CoV-2 IgG antibody levels and binary seropositivity/seronegativity at 6 months as the outcome variable. In reference to the NIBSC standard, the minimum level of anti-SARS-CoV-2 IgG antibodies in baseline samples associated with protection for 6 months was inferred, based on the original dilution of samples.

### Statistical analysis

Data was analysed in Stata 16 (StataCorp. 2019. Stata Statistical Software: Release 16. College Station, TX: StataCorp LLC.) and Graph Pad Prism 9.0 (GraphPad Prism Software, San Diego, California). With respect to demographic data, categorical characteristics were compared using a Chi-Squared test and continuous characteristics compared using the Wilcoxon Rank Sum Test. The distribution of IgG ratios at different time points were compared using the Kolmogorov-Smirnov Test with a false discovery rate approach set at 1% (Benjamini, Krieger and Yekutieli method).

## Results

Following the first wave of the COVID-19 pandemic, the baseline seroprevalence of anti-SARS-CoV-2 spike glycoprotein antibodies in this cohort of DCPs was 16.3% (n=246/1507) (**Table 1**). Ethnicity was a significant risk factor for seropositivity at baseline, with higher seroprevalence observed in individuals of Black ethnicity (35.0%), compared to those of Asian (18.8%) and white ethnicity (14.3%) (p=0.018). There were no differences in seroprevalence between different DCPs; receptionists, who do not have direct patient contact, had the lowest baseline seroprevalence (6.3%), a finding concordant with estimates of seroprevalence in the general population of the West Midlands at the time of baseline sampling [13]. Current smoking was associated with a lower seroprevalence compared to never-smokers or ex-smokers (7.6% vs 16.4% vs 17.6%, p=0.007). Deprivation was associated with a higher seroprevalence: the median index of multiple deprivation rank was 8238 (IQR: 3240, 14408) for seropositive individuals compared to 12081 (IQR: 3858, 21795) for those that were seronegative (p=0.004).

**Table 1:**
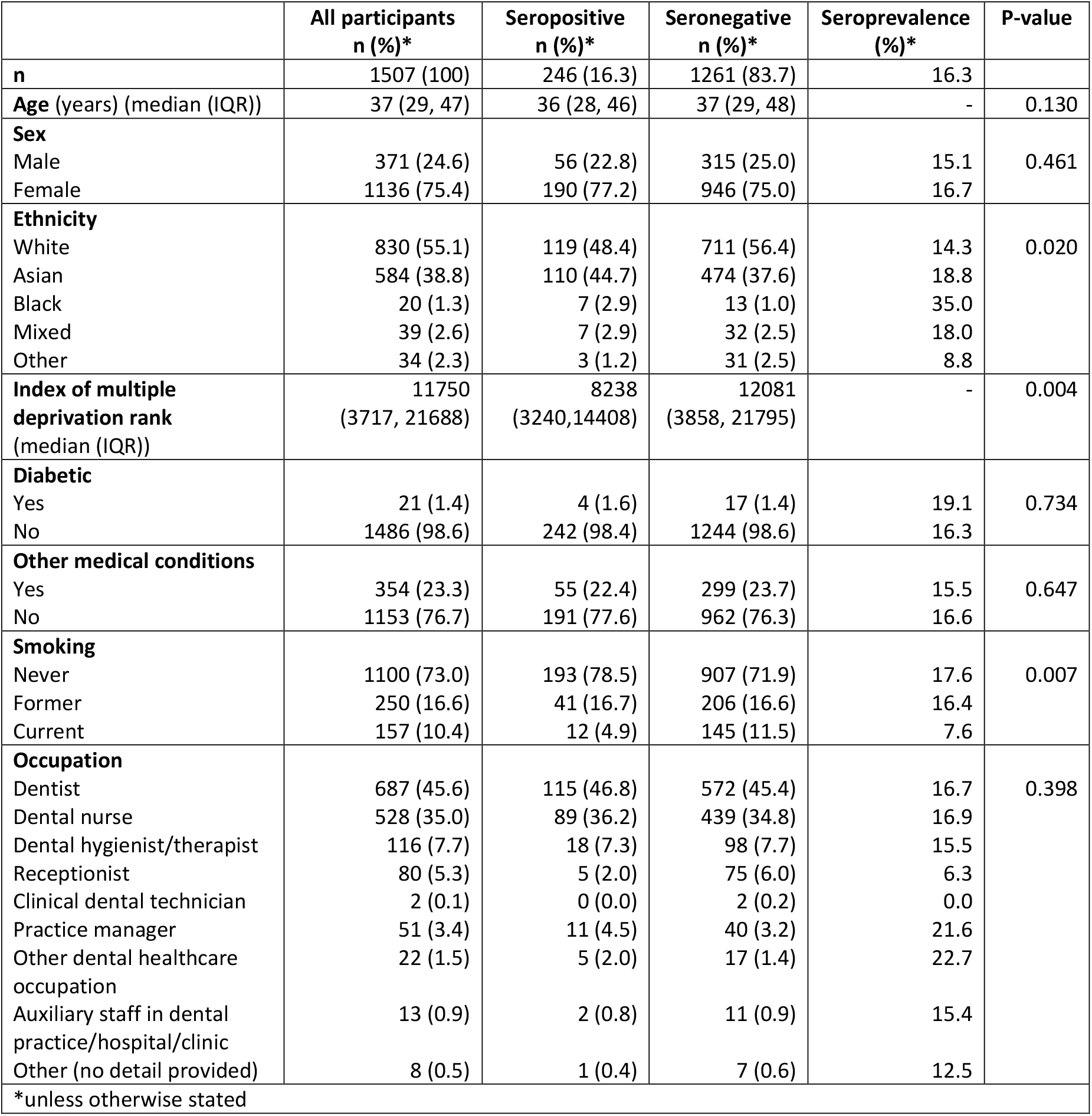
Demographics of study population. Number (%) are presented for categorical and binary characteristics and compared using a Chi-Squared test. Median (interquartile range (IQR)) are presented for continuous characteristics and compared using the Wilcoxon Rank Sum Test.

The cohort was followed longitudinally: individuals who were seropositive at baseline were re-bled at 3 months to study the durability of serological responses (**Figure 1A**). 73% of individuals continued to have a detectable serological response against the spike glycoprotein at three months and, in a subgroup of 79 individuals who were seropositive at baseline and who reattended prior to their vaccination at 6 months, a serological response remained detectable in 72%. Individual IgG and IgA responses were also measured in those who were seropositive. Anti-spike glycoprotein IgG and IgA response were detectable in 42% and 16% of individuals at baseline, reducing to 37% and 9% at 3 months respectively. The discordance between seropositivity defined by the detection of the total antibody response (IgGAM) against the SARS-CoV-2 spike glycoprotein, compared to the IgG isotype, arises from the optimization of the assay for seroepidemiological studies (see Methods).

**Figure 1:**
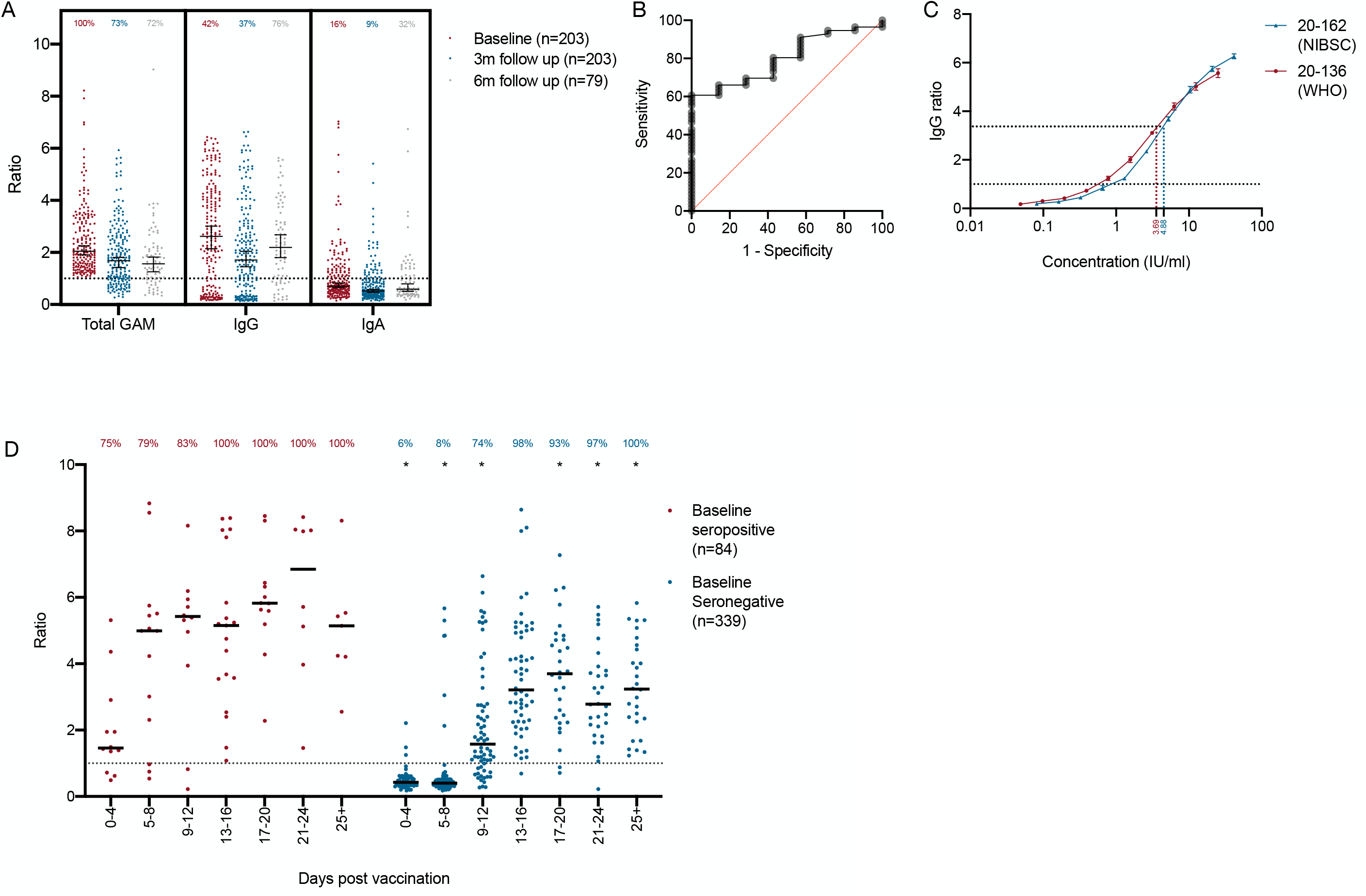
**A** Total anti-SARS-CoV-2 spike glycoprotein antibodies (Total GAM) and IgG and IgA anti SARS-CoV-2 spike glycoprotein antibodies measured at baseline, 3 months and 6 month follow up. At 6 months, only individuals who had not been vaccinated are shown. Data are provided as a ratio of the level of antibody compared to the cutoff calibrator set at 1.0. Percentage of individual above the assay cutoff at each time point is also provided above each column. Median and 95% confidence intervals are shown. **B** Receiver operator characteristic curve describing the relationship between baseline anti SARS-CoV-2 spike glycoprotein IgG ratio and binary IgG seropositivity at 6-month follow up. AUC = 0.80, p=0.01. **C** International reference materials NIBSC 20-136 (WHO) and 20-162 was run in triplicate serial dilutions and the IgG ratio determined. The minimum IgG ratio associated with guaranteed seropositivity 6 months from baseline is shown by the red and blue dotted lines. **D** Kinetics of total antibody response in 423 individuals following a single dose of the Pfizer-BioNTech vaccine. * demonstrate a significant difference (p<0.05) between the distributions of IgGAM ratios at each timepoint following vaccination between individuals who were seropositives and seronegative at baseline as determined by Kolmogorov-Smirnov test. Percentage of individual above the assay cutoff at each time point is also provided.

61.4% (n=926/1507) of the cohort returned questionnaires regarding SARS-CoV-2 infections at 6 months and 59.2% (n=873/1507) were re-bled. In this cohort, 77 PCR-positive SARS-CoV-2 infections were reported by study participants, representing an overall infection risk of 8.3%. The risk of infection was 9.6% in participants who were seronegative at baseline, compared to 2.8% in individuals who were seropositive (p=0.001). As seropositivity at baseline in June 2020 could only be accounted for by SARS-CoV-2 infection, these data suggest that the emergence of antibodies following natural infection with SARS-CoV-2 is associated with a 74% risk reduction for future infection (Risk Ratio 0.26, 95% CI 0.11, 0.63, adjusted for age, sex, ethnicity and smoking).

To further investigate, the phenomenon of re-infection in participants who were seropositive at baseline using the IgGAM assay, participants ‘individual IgG and IgA responses against the SARS-CoV-2 spike glycoprotein reviewed. Reinfections only occurred in the absence of a specific, detectable anti-spike glycoprotein IgG response. Thus, to determine an absolute level of SARS-CoV-2 antibodies associated with ongoing seropositivity at 6 months, we reviewed the baseline anti-spike glycoprotein IgG levels of the 79 participants who had been re-sampled prior to vaccination at 6 months (**Figure 1B**). An IgG ratio greater than 2.97 conferred a likelihood ratio of 2.43 of IgG seropositivity at 6 months (sensitivity 69.7% CI 56.7-80.1%, specificity 71.4% CI 35.9-94.2%); no participant with a baseline IgG ratio greater than 3.36 was IgG seronegative at 6 months (sensitivity 60.7%, CI 47.6-72.4%, specificity 57.0-100.0%). In reference to the first WHO standard for SARS-CoV-2 immunoglobulin (NIBSC 20/136) and the original dilution of the baseline samples, we estimate that the minimum level of anti-SARS-CoV-2 spike glycoprotein IgG antibodies necessary to confer 6 months protection from infection is 147.6 IU/ml (**Figure 1C**). Studies using the NIBSC standard 20/162, generated a similar estimate of 195.2 U/ml.

873 participants were re-bled in January 2021 following 6 months of follow-up. Through natural infection and vaccination, the overall seroprevalence had risen to 49.1% (n=429/873). In individuals who donated serum prior to vaccination and who were seronegative at baseline (n=308), 36 PCR-positive infections occurred and a further 24 asymptomatic seroconversion events had occurred, suggesting the infection risk during the 6-month follow-up period may be as high as 19.4% (n=60/308). 51.5% (n=450/873) had received a single dose of a SARS-CoV-2 vaccine (Oxford/AstraZeneca, n =17; Pfizer-BioNTech, n= 429; Unknown, n=4). The serological responses of individuals receiving a single dose of the Pfizer-BioNTech SARS-CoV-2 were analysed based on prior exposure to the virus - defined by either positive baseline serology, or PCR-proven infection during the follow up period (**Figure 1D**). Vaccination on the background of prior exposure to the virus was associated with a more rapid and quantitatively greater total antibody response against the SARS-CoV-2 spike glycoprotein, consistent with the boosting of immunological memory. In naive participants, greater than 95% seroprevalence was achieved 12 days after vaccination.

## Discussion

Consistent with other studies, we demonstrate that natural infection with SARS-CoV-2 is generally associated with robust and durable serological responses [14, 15]. The percentage of individuals who lose detectable serological responses after 3 months (30%) is higher than that reported in other studies. This is most likely due to the sensitivity of the screening assay that measures the total antibody against the spike glycoprotein [10]. This approach identifies individuals who are exposed to the virus with high sensitivity but some individual appear to mount modest serological responses that are not associated with durable immunity. Nevertheless, in this community based cohort, we corroborate the findings of Lumley et al [5] and Hanrath et al [4] in demonstrating that seropositivity at baseline is associated with protection from infection with SARS-CoV-2. We observed that symptomatic reinfections only occurred in individuals who lacked a robust IgG response, and thus, by relating initial anti-spike glycoprotein IgG levels to the WHO first international reference material for anti-SARS-CoV-2 immunoglobulin (NIBSC 20/136), define a putative antibody level of 147.6 IU/ml that affords a minimum of 6 months protection from reinfection. Clinically, real-world data that relate protection from infection with antibody binding in an in vitro assay is invaluable. Further longitudinal studies in cohorts of individuals following natural infection and vaccination will be necessary to replicate these findings with using assays that employ alternative target SARS-CoV-2 antigens, such as the receptor binding domain, or nucleocapsid.

Vaccination is the most cost-effective and efficacious public health intervention of modern times. In the United Kingdom, the rapid deployment of the Pfizer-BioNTech SARS-CoV-2 vaccination coincided with a planned 6-month follow up of this cohort, affording a unique opportunity to investigate the early serological response to vaccination. Following a single dose of vaccine in naïve recipients, SARS-CoV-2 antibodies were detectable in over 95% of individuals 12 days after vaccination and persisted beyond 25 days post vaccination. In keeping with other contemporaneous studies, we also demonstrate that prior viral infection leads to a more rapid and robust antibody response, consistent secondary immunological responses [16, 17]. The nature and duration of immunity in these cohorts will be critical to understand as the COVID-19 pandemic progresses, particularly with respect to the efficacy of vaccination strategies (single-dose, multiple-doses, vaccine combinations) and in relation to novel viral variants of concern.

Finally, in this community-based cohort of over 1500 individuals, we demonstrate that DCP have a significant occupational risk of exposure to the SARS-CoV-2 virus. The overall baseline seroprevalence in this study of 16.3% exceeds that of the general population in the West Midlands region (6-7%) in June 2020 [13]. The observation that the seroprevalence amongst dental practice receptionists, who have no direct patient contact, was comparable to the general population, supports the hypothesis that occupational risk arose from close exposure to patients. Seroprevalence across the West Midlands region increased by 12.3% between June 2020 and January 2021 [18]; the risk of PCR-proven infection in seronegative DCPs in our study during this time was 11.7%. Further studies are necessary to comprehensively understand whether these comparative statistics represent a lowering of exposure rates of DCPs to background population levels following reopening of general dental practices and the additional precautions taken to ensure practices became COVID-19 secure.

## Data Availability

Data are available for a period of 24 months from publication upon reasonable request. Proposals should be directed to the corresponding authors.

## Acknowledgements

The authors are grateful to the staff of the University of Birmingham Clinical Immunology Service for facilitating laboratory studies. The authors are also grateful to Joanne Crumpler, Jacqui Rees, Joanne Whitehouse, Joanna Rooney, Amneet Sidhu, Amanda Stokes, Harpreet Dhami, Marie Jones, Kate Ward, Erika Malone, Jessica Chapple and Yvonne Simaloi for their help supporting this study. Finally, the authors are indebted to the DCPs in the West Midlands region who volunteered to participate in this study.

## Ethical approval

The study was approved by the London - Camden and Kings Cross Research Ethics Committee (reference 20/HRA/1817). All participants provided written informed consent prior to enrolment in the study

## Abbreviations

AGP: Aerosol generating procedures
COVID-19: Coronavirus disease 2019
DCP: Dental care professionals
GDP: General dental practitioners
IRM: International reference material
NIBSC: National Institute for Biological Standards and Control
PCR: Polymerase chain reaction
SARS-CoV-2: Severe acute respiratory syndrome coronavirus 2
WHO: World Health Organization

